# Protein misfolding in the gastrointestinal tract predicts and prognosticates neurodegenerative disease years before symptom onset

**DOI:** 10.1101/2025.10.13.25337884

**Authors:** Tatiana Langerová, Emma MacVicar, Rollo Press, Fiona L. Read, Mattia Piana, Sarah Murdoch, Jayne Innes, Joan Wilson, Angus J. M. Watson, Fergal M. Waldron, George Ramsay, Jenna M. Gregory

**Affiliations:** Institute of Medical Sciences, University of Aberdeen, Foresterhill, Aberdeen, UK; Aberdeen Centre for Evaluation, University of Aberdeen, Foresterhill, Aberdeen, UK; Department of Colorectal Surgery, Aberdeen Royal Infirmary, Foresterhill, Aberdeen, UK; School of Medicine, Medical Sciences, and Nutrition, Foresterhill, Aberdeen, UK; NHS Grampian Biorepository, Department of Pathology, Aberdeen Royal Infirmary, Foresterhill, Aberdeen, UK; Institute of Applied Health Sciences, University of Aberdeen, Foresterhill, Aberdeen, UK; Department of Colorectal Surgery, Raigmore Hospital, Inverness, UK

**Author notes:** Correspondence to, 01224438063. Correspondence to, 01224 437524. Equal contributions (Tatiana Langerová, Emma MacVicar). Equal contributions (Fergal M. Waldron, George Ramsay, Jenna M. Gregory). Lead contact: Jenna Gregory (, 01224 437524).

**Keywords:** Neurodegeneration, Proteinopathy, TDP-43, Alpha-synuclein, Tau, Gastrointestinal tract, Biomarker

## Abstract

**Background:** Disease-modifying therapies for neurodegenerative disorders are unlikely to succeed once symptoms emerge, as significant neuronal loss has already occurred. Accessible biomarkers that predict disease years in advance, and that can serve as target-engagement readouts for prevention trials, are urgently needed.

**Methods:** We analysed archival gastrointestinal (GI) biopsies from 196 individuals with unexplained GI symptoms and 13–15 years of follow-up. Using sensitive histopathological staining, we assessed misfolded TDP-43, tau, and α-synuclein to test whether peripheral proteinopathies can serve as predictive biomarkers for neurodegeneration.

**Results:** Protein misfolding enteropathy was detected in 60% of cases. Individuals with GI proteinopathy were significantly more likely to develop non-Alzheimer’s dementia or α-synucleinopathies, with >80% sensitivity. The presence of two or more proteinopathy markers was associated with a dose-dependent reduction in survival, establishing GI proteinopathy as an independent, life-limiting prognostic factor. Importantly, these pathological changes were present 6.9 years before neurological symptoms emerged.

**Interpretation:** Our findings reveal that neurodegeneration-associated proteinopathies are not confined to the central nervous system but can be detected in routine GI biopsies years before clinical onset. This discovery provides a practical and scalable biomarker platform that could transform early diagnosis, risk stratification, and target-engagement monitoring in clinical trials. Protein misfolding enteropathy represents a new frontier for disease interception in neurodegenerative disorders, enabling intervention at a stage when neuronal damage may still be preventable.

**Funding:** Target ALS, LifeArc, NHS Grampian 45

**Graphical Abstract:** 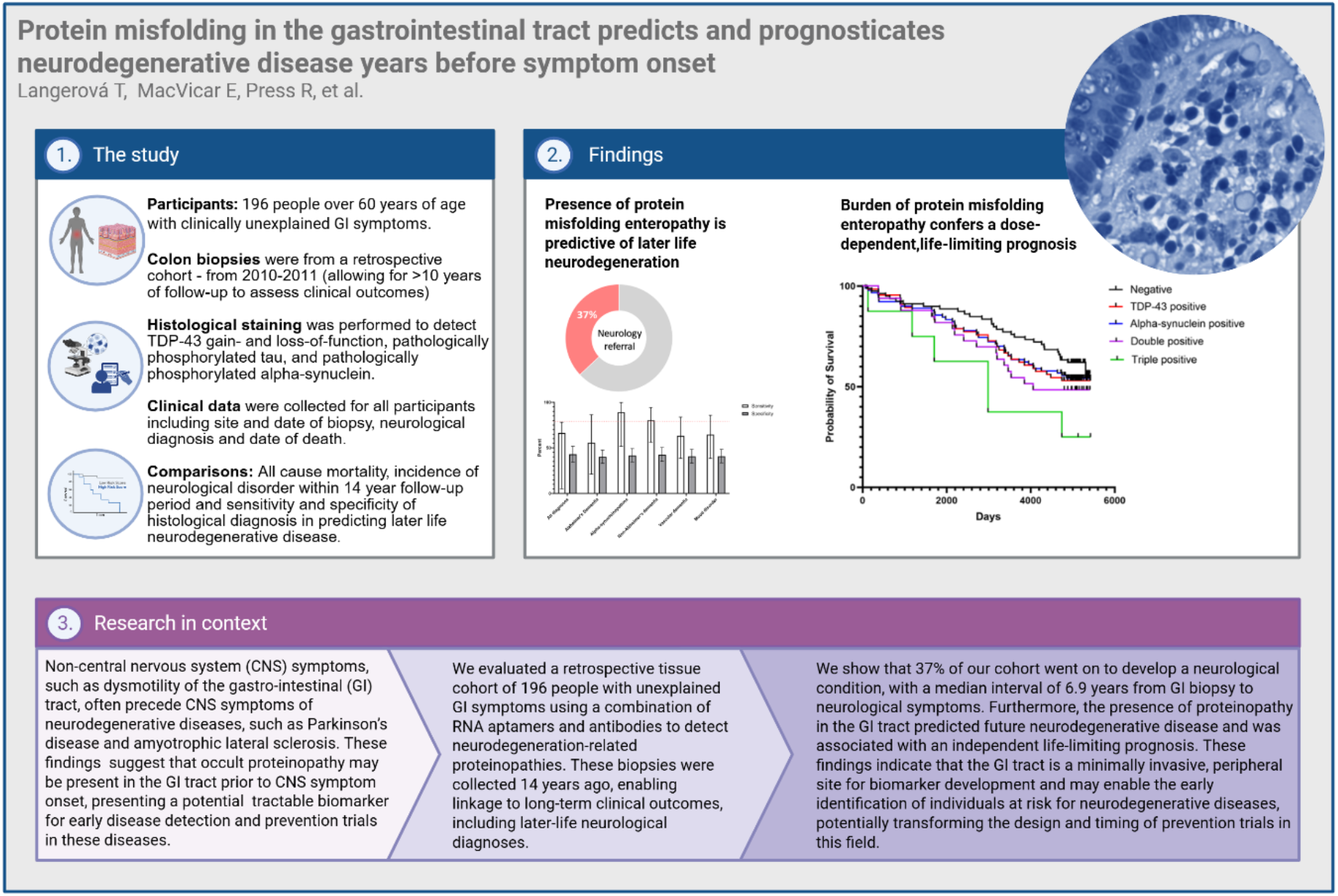

**What is already known on this topic:** Neurodegenerative diseases are typically diagnosed after symptoms emerge, by which time extensive and irreversible neuronal loss has occurred. Reliable biomarkers that can identify individuals at risk many years earlier are lacking, and current strategies for early detection and monitoring of disease-modifying therapies remain limited.

**What this study adds?:** This study shows that protein misfolding enteropathy, detectable in routine gastrointestinal biopsies, predicts the later development of non-Alzheimer’s dementia and α-synucleinopathies with high sensitivity. Pathological protein deposits were present more than a decade before neurological symptoms, and the presence of multiple proteinopathy markers correlated with reduced survival.

**How this study might affect research, practice or policy:** These findings establish gastrointestinal proteinopathy as a practical, scalable biomarker for early detection and prognosis in neurodegenerative disorders. This approach could enable earlier intervention, improve risk stratification, and provide target-engagement readouts for clinical trials, opening new avenues for disease interception strategies.

## Introduction

Accumulation of misfolded proteins in the central nervous system (CNS) is a common pathological hallmark seen in many neurodegenerative diseases including amyotrophic lateral sclerosis (ALS), Alzheimer’s disease (AD), Parkinson’s disease (PD), and cognitive ageing.^1^ However, at the time of symptom onset, there is already substantial neuronal cell loss with irreparable damage done to the CNS.^2^ Intervention at the point of symptom onset may therefore be too late resulting in a relative lack of translational progress, minimal treatment options, and no cure. Early intervention strategies aligned with readily accessible targets for biomarker development are urgently needed for these diseases. Indeed, non-CNS symptoms, such as dysmotility of the gastro-intestinal (GI) tract, often precede CNS symptoms of ALS, PD, and AD.^3-6^ In line with this, misfolded proteins associated with these diseases such as TDP-43, alpha-synuclein, and tau are not restricted to the CNS, occurring in other organ systems, including the gut.^7-9^ Furthermore, a recent population study, evaluating over 1 million people, demonstrated that people over the age of 60 with unresolved GI symptoms that had a diagnostic colon biopsy, reported as histologically normal, had an increased risk of later life ALS (HR = 1.24; 95% CI 1.04 to 1.42).^10^ This is further supported by the peripheral identification of early markers of pathological TDP-43 loss-of-function, such as TDP-43-dependent cryptic exons, that can be detected nearly a decade prior to the presence of TDP-43 cytoplasmic aggregates in people with AD and cognitive ageing.^11^ Taken together, these studies imply that we may be missing occult, symptomatic, systemic proteinopathies long before CNS symptoms begin.

Expanding upon this work, with the aim of developing an early predictive biomarker for neurodegenerative diseases caused by protein misfolding, we focused on the GI tract as a readily accessible peripheral site for biopsy and tissue diagnosis. We hypothesised that individuals over the age of 60, with histologically unexplained GI symptoms represented a high-risk population^10^ that could be targeted for future precision prevention strategies aimed at reducing systemic protein misfolding pathology and later life risk of neurodegeneration. To test this hypothesis, we have built a retrospective cohort of people over 60 years of age, who had GI symptoms with a histologically normal diagnosis. The cohort comprised investigative biopsies taken from 196 individuals, who fit these criteria, in 2010-2011 with over 10 years of intervening clinical follow-up data. Using these formalin-fixed, paraffin embedded, archival biopsies, we implemented an immunohistochemical staining strategy employing sensitive staining techniques to identify disease-associated pathological alpha-synuclein, tau, and TDP-43. We also implemented a novel detection strategy (an RNA-aptamer that detects pathological TDP-43, called TDP-43^APT^) for identifying TDP-43 pathology with improved sensitivity compared to antibody-based approaches^12,13^ and a cryptic exon *in situ* hybridisation probe to demonstrate TDP-43 loss-of-function.^12^ We then performed a clinicopathological comparison to determine the ability of these histological stains to predict future neurodegenerative disease risk and survival.

## Methods

### Case selection, data extraction, and ethics approvals

This study uses archival biopsies acquired from the NHS Grampian Biorepository under existing ethical frameworks: Research type: Research Tissue Bank; IRAS ID: 296502; REC name: North of Scotland Research Ethics Committee 2; REC reference: 21-NS-0047. Tissue samples were requested from the NHS Grampian Biorepository, under existing ethical frameworks with a tissue request number of TR301. The requirement for written informed consent is waived for archival tissue samples, ethics covers the use of unconsented material provided it is anonymised at point of availability to the researchers.

Colonic biopsy material was requested from individuals that have undergone a diagnostic colonoscopy, but whose biopsy was determined to not contain any histological anomalies. Formalin-fixed-paraffin-embedded (FFPE) tissue samples were collected from an “initial tissue request cohort” of 210 individuals (figure 1A). As illustrated in figure 1B, from this initial cohort of 210, four individuals had to be excluded from the study as they did not fit all study criteria, with a further ten individuals excluded due to a lack of sufficient clinical follow-up data. This left a final “study cohort” of 196 individuals for whom tissue biopsies and clinical follow-up data were available.

**Figure 1.**
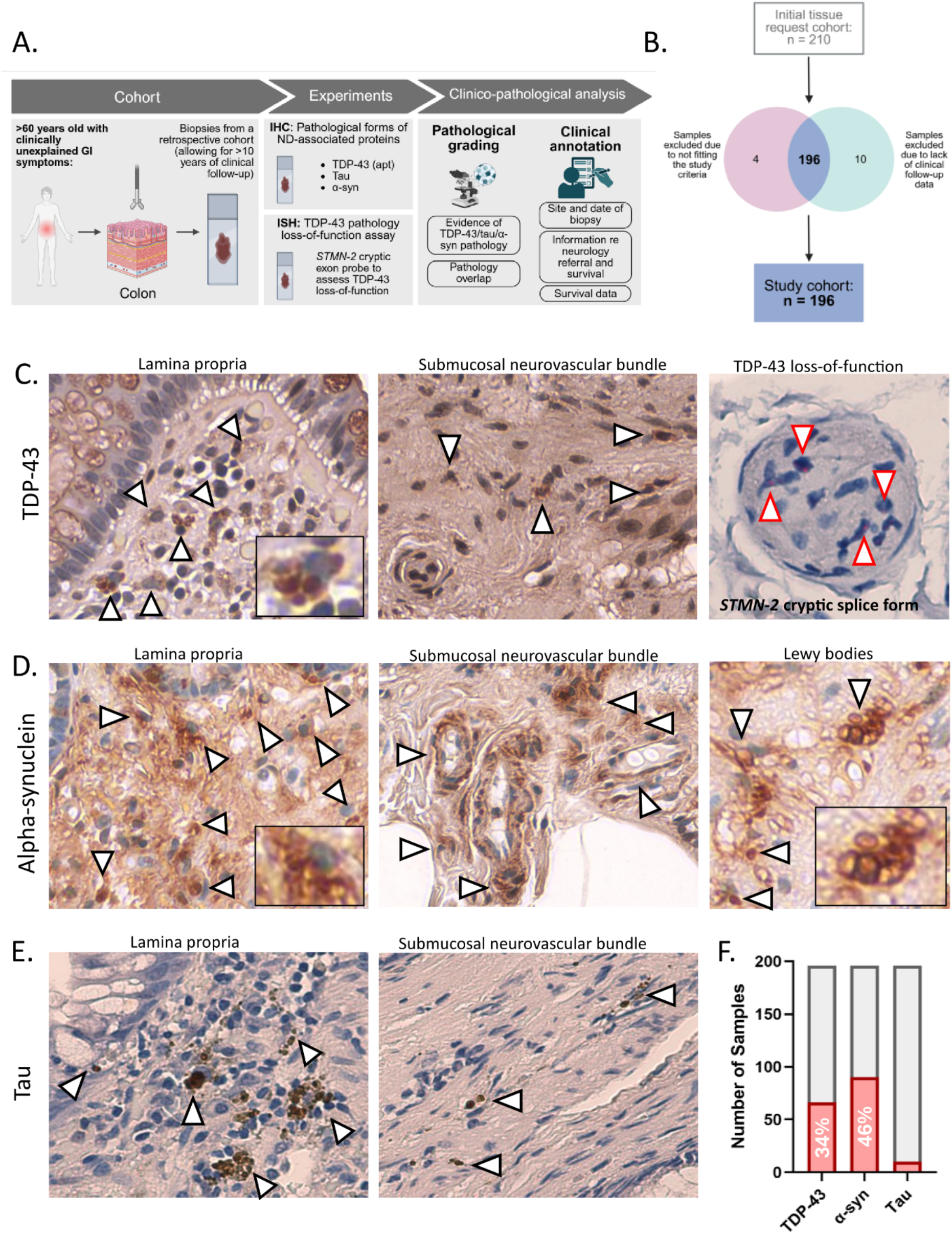
Occult protein misfolding pathology is common in individuals presenting with histologically unexplained gastrointestinal symptoms. **A**. Experimental workflow including details regarding the retrospective study cohort, experimental details, and data collected for clinico-pathological investigation. **B**. Illustration of the study cohort assembly methodology. Formalin-fixed-paraffin-embedded (FFPE) tissue samples were collected from an “initial tissue request cohort” of 210 individuals. From this initial cohort of 210, four individuals were excluded from the study as they did not fit all study criteria, with a further ten individuals excluded due to a lack of sufficient clinical follow-up data. This left a final “study cohort” of 196 individuals for whom tissue biopsies and clinical follow-up data were available **C**. Representative photomicrographs taken from a case with TDP-43 pathology, detected using TDP-43 RNA aptamer (TDP-43^APT^). Left image demonstrates pathological intracellular TDP-43 aggregates (white arrowheads) within the lamina propria of the colon. Inset box is an optical zoom of the cell indicated by a black arrowhead. Scale bar = 20 μm. Centre image demonstrates pathological intracellular TDP-43 aggregates (white arrowheads) within the submucosal neurovascular bundle of the colon. Scale bar = 20 μm. Right image is a cross-cut submucosal nerve bundle in the colon, within which there is demonstrable evidence of individual mRNA molecules of *STMN-2* that contain a cryptic exon (single red dots) that would not be there if TDP-43 was functioning correctly. This *in situ* hybridisation technique, using BaseScope™ probes, is specifically designed to bind to the pathological cryptic exon demonstrating TDP-43 loss-of-function. Scale bar = 10 μm. **D**. Representative photomicrographs taken from a case with aggregation of a disease-specific phosphorylated form of alpha-synuclein (phospho-alpha-syn). Left image demonstrates pathological intracellular alpha-synuclein aggregates (white arrowheads) within the lamina propria of the colon. Inset box is an optical zoom of the cell indicated by a black arrowhead. Scale bar = 20 μm. Centre image demonstrates pathological intracellular alpha-synuclein aggregates (white arrowheads) within the submucosal neurovascular bundle of the colon. Scale bar = 20 μm. Right image demonstrates Lewy bodies (white arrowheads) detectable within both the lamina propria and submucosal tissues of affected cases. Inset box is an optical zoom of the region indicated by a black arrowhead. Scale bar = 10 μm. **E**. Representative photomicrographs taken from a case with aggregation of a disease-specific phosphorylated form of tau (using the AT8 antibody). Image demonstrates pathological intracellular tau aggregates (white arrowheads) within the lamina propria of the colon. **F**. Frequency distribution indicating the number of cases that were pathologically graded as positive for each proteinopathy.

Clinical data were collected by clinicians (EM, RP, AJMW, and GR) according to the Caldicott Principles. The NHS Grampian Biorepository team (MP, SM, JI, JW) maintained the data on a secure server providing only anonymised, non-identifiable, clinical information to the research team. The principles were agreed to by the team in advance of the work being carried out and documented in the tissue request form. All research investigators involved with tissue staining (TL, FLR, FMW, and JMG) were kept anonymised to all clinical and demographic data. Biopsies were retrieved from 2009-2011 to allow for between 13-15 years (median 14.03; IQR 12.92 to 14.44) of follow-up data to determine whether an individual had gone on to develop neurological symptoms. Anonymised data (using study IDs provided by the Biorepository) were used to plot (i) survival (time from tissue biopsy to date of death), (ii) sensitivity and specificity for detecting a later life diagnosis, and (iii) the site of biopsy.

The study cohort of 196 individuals was balanced for sex (χ^2^=0.735, p= 0.391) with 104 (53.1%) females and 92 (46.9%) males. The median age at biopsy was 70 (IQR 67 to 72) years, with age ranging from 64-76 years in the cohort. Biopsies were taken over a 2-year period from December 2009 until December 2011. To allow for a minimum >10-year follow-up period, post-biopsy follow-up survival data were retrieved over a 14-year period, from the earliest death recorded in September 2010 to the most recent death in November 2024. The number of people who had died over the study period was 81 (41.32% of the total 196), with 115 (58.67% of 196) individuals still alive on 11/11/24, the date of the study end. Detailed study cohort characteristics are provided in Table 1. 134

**Table 1.**
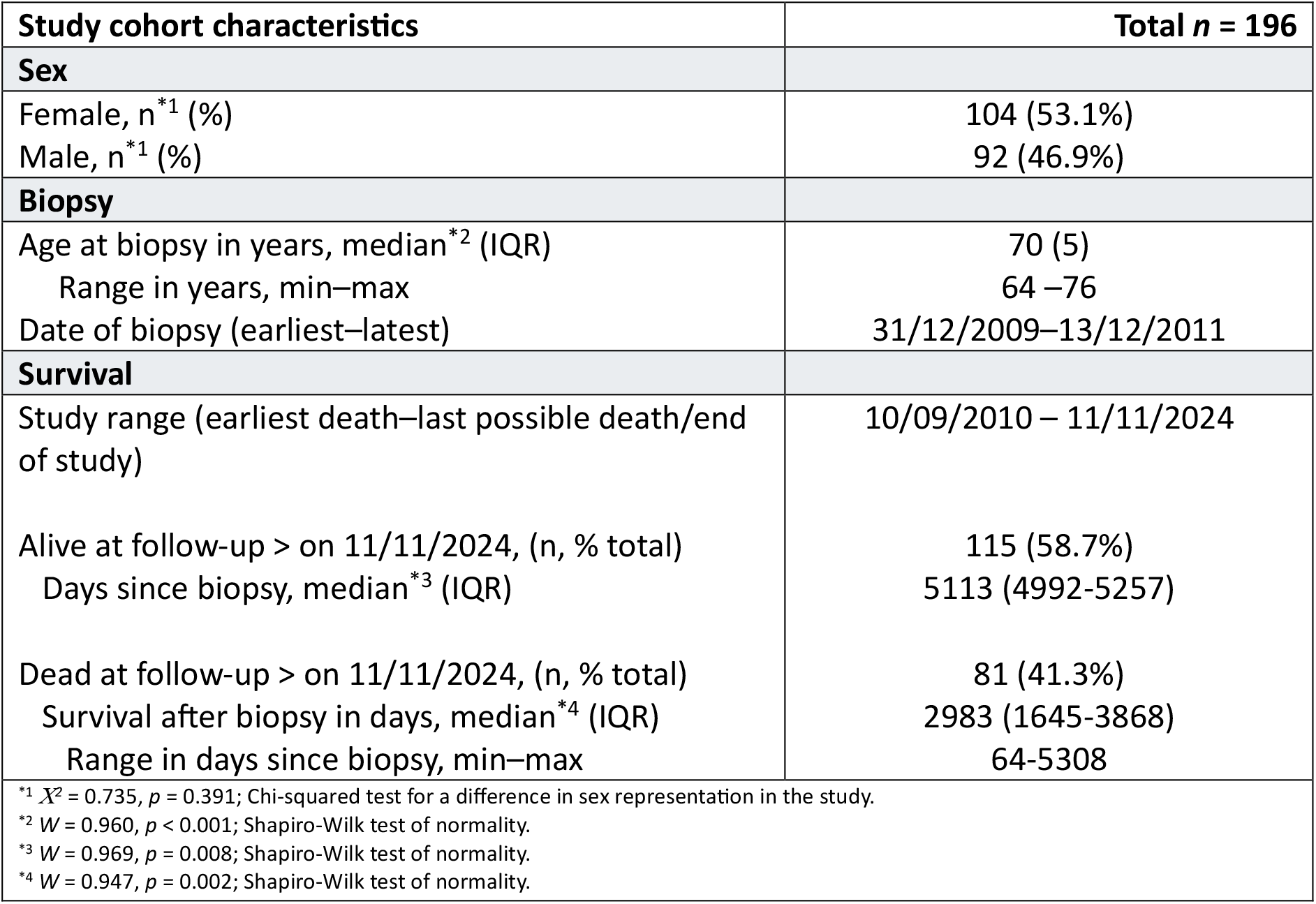
Study cohort characteristics. Study cohort characteristics table with information on patient sex, age at biopsy, date of biopsy and survival over the course of the study. Test statistics with p-values are provided for; tests for sex bias in the cohort, and normality of data to justify the reporting of medians with interquartile ranges where appropriate.

### Immunohistochemistry

To assess the presence of pathological protein inclusions in the archival biopsy tissue of our retrospective cohort, standard immunohistochemistry (IHC) and TDP-43 RNA aptamer staining protocols were performed on the “tissue cohort”, as published previously.^12^ Target antigen retrieval was performed using citric acid at pH 6 in a pressure cooker. TDP-43 aptamer (TDP-43^APT^ CGGUGUUGCU with a 3’ Biotin-TEG modification, ATDBio, Southampton, UK) was diluted in distilled water to a final concentration of 156 nM. Phospho-α-Syn (AB59264, Abcam) and anti-tau (AT8; MN1020, Thermo Fisher Scientific) antibodies were diluted in TBS 1:75 and 1:2000 respectively. Antibodies were incubated for one hour at room temperature for the phospho-α-Syn antibody and 30 minutes at 4°C for the AT8 antibody.

### In situ *hybridisation*

To evaluate TDP-43 loss-of-function, a BaseScope™ *in situ* hybridisation probe was used to detect pathological cryptic exon containing *Stathmin-2* (*STMN-2*) mRNA transcripts. STMN-2 is a neuronal protein whose splicing is mediated by TDP-43. If TDP-43 malfunctions, it fails to inhibit the integration of cryptic exons into the mature protein, resulting in the formation of truncated STMN-2 containing an aberrant cryptic exon. Detection of *STMN-2* cryptic exons thus indicates TDP-43 loss-of-function. Our previously published BaseScope™ protocol was performed without additional alterations^12^ using our *STMN-2* cryptic exon probe (STMN-2(CE); Cat. no. 1048241-C1, ACD-Bio). 153

### Statistics

Confidence intervals of proteinopathy incidences and dementia diagnosis rates were computed using the modified *Wald* method of Agresti and Coull^14^ implemented in the binom R package.^15^ The Agresti-Caffo^16^ method was used to compute confidence intervals for the difference between two proportions, and to test for a difference between them. Hypothesis tests for differences in median ages of biopsy across instances of proteinopathies (none, single, double, triple) were carried out using Kruskal-Wallis test. Time-to-event analysis are presented as Kaplan-Meier curves, with log-rank (Mantel-Cox) tests for differences between curves. Sensitivity and specificity for predicting later life risk of neurological diagnosis was carried out in GraphPad Prism version 10.0.0 for Windows, GraphPad Software, Boston, Massachusetts USA, www.graphpad.com or R (version 4-5.1).

## Results

### Incidence: GI biopsies harbour misfolded proteins, even in people with “normal” histology

To establish whether the GI tract could be exploited as a readily accessible site for predictive/target engagement biomarker development in neurodegenerative diseases, we first established an investigative retrospective cohort of archival biopsy material. Learning from previous findings that people with histologically unexplained GI symptoms, have an increased risk of ALS^10^ we used these criteria to develop our own cohort. We retrieved the colon biopsies our retrospective cohort and performed immunohistochemical staining for three neurodegenerative disease-associated markers, selected for their specificity to detect pathological forms of these proteins only. We used an RNA aptamer to stain TDP-43 (TDP-43^APT^), AT8 anti-tau antibody to detect pathologically phosphorylated tau, and the phospho-S129 antibody to detect pathologically phosphorylated alpha-synuclein. We also used an *in-situ* hybridisation probe designed to detect TDP-43 loss-of-function dependent inclusion of a cryptic exon in *STMN-2* mRNA transcripts (figure 1A).

Using this multimodal histopathological approach in 196 individuals (figure 1B), we detected pathological TDP-43 (figure 1C), α-synuclein (figure 1D), and tau (figure 1E) in archival colonic biopsies from individuals with unexplained gastrointestinal symptoms. Aggregates were observed in both the lamina propria and submucosal neurovascular bundles, with evidence of TDP-43 loss-of-function in enteric nerve bundles (figure 1C). Overall, 34% of individuals harboured TDP-43 pathology (66/196; 95% CI 27.4–40.6), 46% harboured α-synuclein pathology (90/196; 95% CI 39.1–52.9), and 5% harboured tau pathology (10/196; 95% CI 2.7–9.3) (figure 1F). Tau deposition occurred exclusively in cases with multiple pathologies. No sex differences were observed in the prevalence of TDP-43 (χ^2^=0.583, p=0.445), alpha-synuclein (χ^2^= 0.005, p=0.942), or tau (χ^2^=0.016, p = 0.900) pathologies. These data demonstrate that protein misfolding enteropathy (PME) is common and detectable in routine GI biopsies, even in cases deemed histologically normal at the time of sampling. 188

### Prediction: PME predicts specific types of neurodegeneration with high sensitivity

Clinical follow-up data were available for all 196 individuals, with a median of 14 years (IQR 12.9–14.4) after biopsy. During this period, 37% (73/196; 95% CI 31.1–44.4) were referred to a neurological clinic for new symptoms (figure 2A). The likelihood of referral did not differ by sex (χ^2^=0.34, p=0.56).

**Figure 2.**
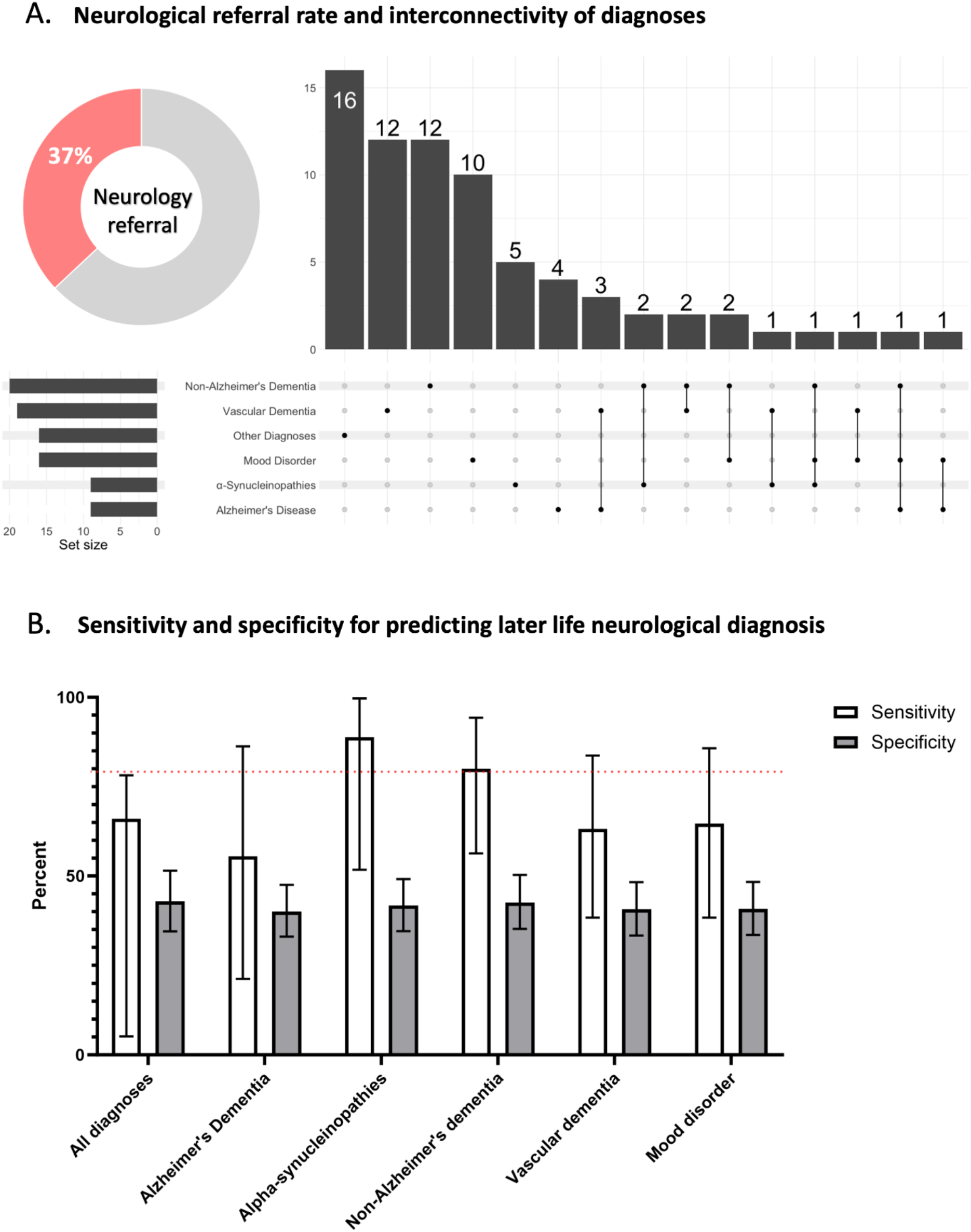
Protein misfolding enteropathy is a sensitive predictor of later life risk of non-Alzheimer’s dementia and alpha-synucleinopathies. **A**. Donut chart indicating the percentage of all cases that went on to develop neurological or neuropsychiatric symptoms of neurodegeneration and receive a neurological diagnosis within the clinical follow-up period of 13-15 years (median 14.03; IQR 12.92 to 14.44) adjacent to an upset plot demonstrating interconnectivity of diagnosis frequency within the cohort. **B**. Graphical depiction of sensitivity and specificity (with 95% confidence intervals) calculated for each of the diagnostic categories. Sensitivity in white and specificity in grey. Red line indicates an 80% cut off below which diagnostic accuracy would be insufficient to warrant further investigation. Specificity of the test (biopsy staining for GI tract proteinopathy) is poor, consistently below 50%. This indicates that if you have no evidence of GI tract proteinopathy, you may still be at an increased risk of neurodegeneration (high false negative rate). However, specificity is high (above 80%) for non-Alzheimer’s dementias (e.g. frontotemporal dementia) and for alpha-synucleinopathies (e.g. Parkinson’s disease or Ley body dementia) – indicating that if you have GI tract proteinopathy you develop symptoms for these diseases within 15 years (high true positive rate).

Baseline evidence of gastrointestinal protein misfolding enteropathy (PME) was strongly associated with later development of neurodegenerative disease (figure 2B). PME predicted non-Alzheimer’s dementia and α-synucleinopathies (including Parkinson’s disease and Lewy body dementia) with >80% sensitivity. In contrast, predictive value was poor for Alzheimer’s dementia, vascular dementia, and new-onset mood disorders. Specificity was modest across all diagnostic categories (<50%), indicating that absence of PME does not exclude future neurological disease (figure 2B).

These findings establish PME as a selective, sensitive biomarker for dementias driven by TDP-43 and α-synuclein pathology, while highlighting its limited relevance for Alzheimer’s and vascular dementias. Importantly, this predictive signal was detectable more than a decade before the onset of neurological symptoms, supporting gastrointestinal biopsy as a practical, early-access biomarker platform for neurodegeneration.

### Co-pathology: Multiple pathologies co-occur, reflecting CNS disease and supporting PME as a mechanistic biomarker

Having investigated the incidence of individual proteinopathies found in the gut, we wanted to know how commonly these proteinopathies co-occur. No TDP-43, alpha-synuclein or tau proteinopathy could be detected in 40% (78/196; 95% CI 33.2 to 46.78) of individuals in the study cohort. Single proteinopathies were identified at a similar incidence of 39% (77/196; 95% CI 32.71 to 46.27) with double proteinopathies in 17% (33/196; 95% CI to 12.21 to 22.74) of individuals, and triple proteinopathies in 4% (8/196; 95% CI to 1.95 to 7.98). Overall, a single, double or triple proteinopathy was detected in 60% (118/196; 95% CI 53.22 to 66.80) of individuals – therefore, the ratio of no proteinopathies, to single proteinopathies, to multiple proteinopathies was approximately 40:40:20 (see figure 3A). There was no evidence for an effect of age at time of biopsy (χ^2^=2.304, p=0.512), or sex differences (χ^2^=1.175, p=0.759) on the presence/absence, or number of proteinopathies (figure 3E).

**Figure 3.**
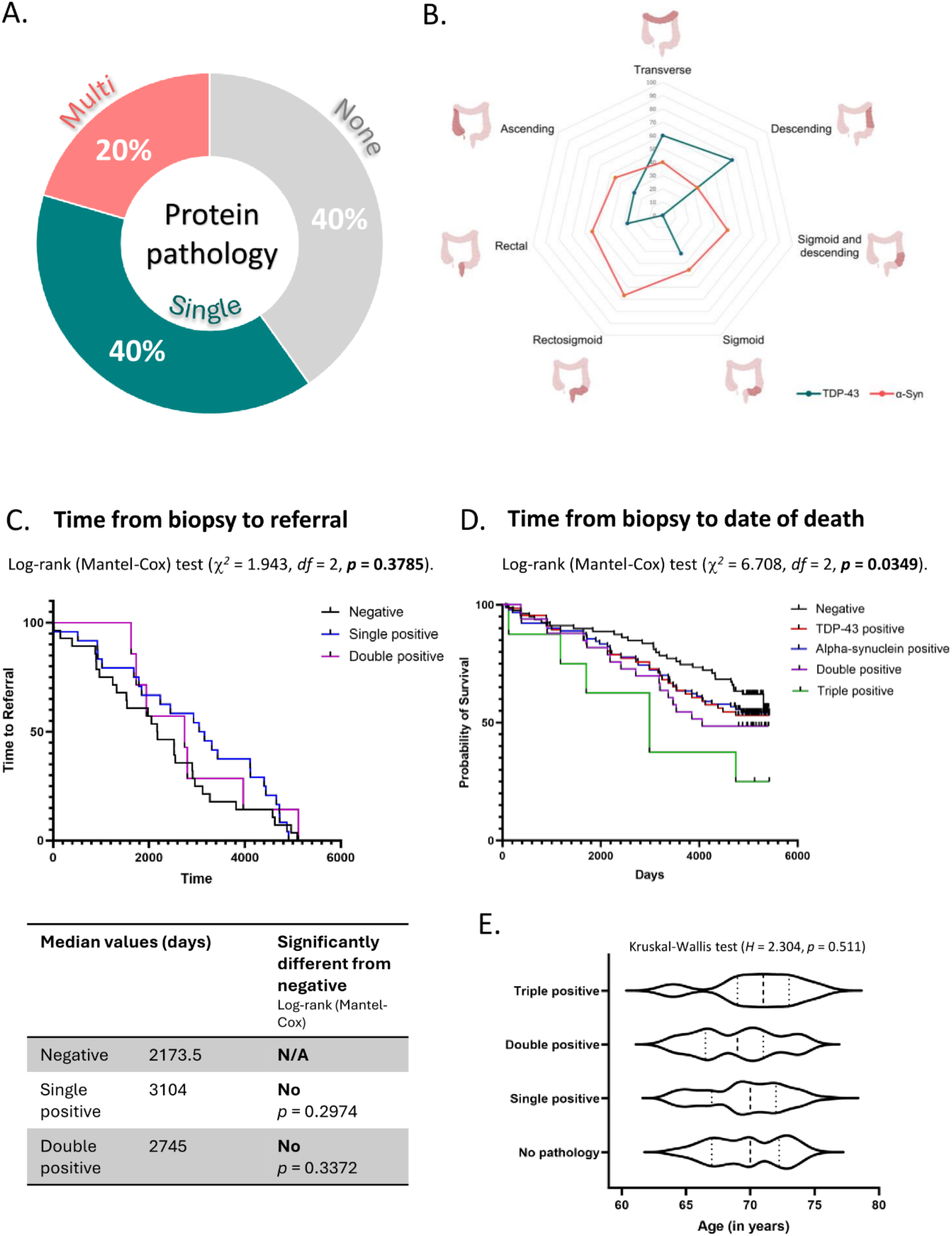
Protein misfolding enteropathy confers a life limiting prognosis. **A**. Donut chart indicating the percentage of the cohort that demonstrated single (either alpha-synuclein or TDP-43 pathology) or multi-proteinopathy (two or more of tau, alpha-synuclein, and TDP-43). Tau was only ever seen in the context of multi-proteinopathy and was never observed as a single proteinopathy. **B**. Radar plot demonstrating the distribution of pathology within the lower GI tract for alpha-synuclein and TDP-43 (too few cases were observed to include tau). Despite multi-proteinopathy being commonly observed for these stains, the distribution was distinct but overlapping. TDP-43 pathology was predominantly in the transverse and descending colon (green line) and alpha-synuclein in the distal colon including the sigmoid and rectum (red line). **C**. Kaplan-Meier plot demonstrating time-to-event analysis from date of biopsy to date of neurological diagnosis. The median time to diagnosis was 6.9 years and was not statistically significant between groups based on presence or extent of proteinopathy. Median time-to-event data are included in the table below the plot. **D**. Kaplan-Meier survival curve indicating a significant dose-dependent effect of multi-proteinopathy on survival across the 13–15 year (median 14.03; IQR 12.92 to 14.44) follow-up period. Survival curves for negative, double, and triple (demonstrating dose-dependence) are significantly different using log-rank (Mantel-Cox) test (χ^2^=6.708, df=2, p=0.0349). **E**. Violin plot demonstrating no difference between groups for age of biopsy, demonstrating that age was not a confounding factor in the survival analysis in D.

We noted that many cases demonstrated evidence of double (usually TDP-43 and alpha-synuclein) and triple positivity. Tau pathology did not occur outside of the context of a multi-proteinopathy, whereas TDP-43 and alpha-synuclein were observed as both single and double proteinopathies (figure 3A). Due to this overlap, 60% of cases within our cohort had at least one detectable form of proteinopathy. Despite TDP-43 and alpha-synuclein frequently co-occurring in this cohort, their anatomical distribution in the gastrointestinal tract was distinct (figure 3B). TDP-43 was more frequently observed in biopsies from the transverse and descending colon, whereas alpha-synuclein pathology was more frequently observed in the sigmoid colon and rectum. Time from biopsy to neurological referral was consistent across the cohort (median time to diagnosis = 2527 days or 6.9 years) and was not dependent (χ^2^=1.943, p=0.379), on the presence or extent of protein enteropathy (figure 3C). 227

### Prognosis: PME is life-limiting, justifying surveillance and prevention strategies

Finally, we wanted to test whether PME has any effect on patient outcomes following biopsy. This was an important question to ask as this cohort of individuals will have been, prior to these findings, discharged from tertiary care services with a normal colonoscopy and normal histology. They will therefore not have received any further treatment or follow-up, despite having an undetected proteinopathy. Survival analysis was performed from date of biopsy to date of death for those that were deceased. Those that were still alive at time of data collection were censored at the end point of data collection.

Survival analysis revealed a dose-dependent effect: individuals with double or triple PME had significantly reduced survival compared to those without PME. Importantly, these individuals had been discharged with “normal” histology, underscoring that PME represents a clinically silent but prognostically significant condition. Thus, routine GI biopsy not only predicts future neurodegeneration but also identifies a life-limiting disorder previously overlooked in standard care.

## Discussion

In this study, we describe a life-limiting pathological entity − which we call *protein misfolding enteropathy* − found in the GI tract of people over the age of 60 who have GI symptoms of dysmotility. The investigative cohort that allowed this discovery consists of individuals aged over 60 who have had no histological abnormality noted on their diagnostic colon biopsies, despite their adverse GI symptoms. Using an archival tissue cohort (n=196), we performed immunohistochemical staining detecting three distinct markers of neurodegenerative diseases (pathological forms of TDP-43, alpha-synuclein, and tau). We show that TDP-43 aggregates are present in 34% (66/196; 95% CI 27.42 to 40.55) of individuals, alpha-synuclein aggregates in 46% (90/196; 95% CI 39.09 to 52.91), and tau aggregates in 5% (10/196; 95% CI 2.68 to 9.25). Indeed, accounting for cases with co-pathology, some form of proteinopathy is found in 60% (118/196; 95% CI 53.22 to 66.80) of individuals in this cohort – individuals who are being sent home, reassured that there have been no abnormalities identified on colonoscopy or pathological examination, however, retrospectively have occult proteinopathy that may have explained their symptoms. Indeed, we know that alpha-synuclein pathology in the gut can cause neuronal loss and corresponding GI symptoms, including constipation.^17^ Furthermore, robust epidemiological data exists linking dysmotility and a wide range of neurodegenerative diseases^18,19^, a possible mechanism for which, i.e. *protein misfolding enteropathy*, we report here.

Our retrospective cohort (n=196) allowed for 13-15 (median 14.03; IQR 12.92 to 14.44) years of clinical follow-up, following GI symptom onset. During this period 37% (73/196; 95% CI 31.08 to 44.39), displayed neurological symptoms warranting a referral to secondary care services. Because the rate of neurological symptom referral within this cohort was similar to the rate of TDP-43 and alpha-synuclein pathologies found in gut samples, we next explored the predictive power of GI proteinopathies in detecting a later life onset of neurological disease. While the specificity was low, sensitivity was over 80% for detecting non-Alzheimer’s dementias (such as ALS-FTSD) and alpha-synucleinopathies (such as PD and Lewy body dementia), which suggests that detectable protein misfolding enteropathy is associated with an increased risk of these conditions, corroborating the epidemiology data reported previously.^10^ We posit that the presence of *protein misfolding enteropathy* represents a potential biomarker for certain neurodegenerative diseases. Currently, other diagnostic tests with predictive sensitivity of 80% are routinely used in clinic such as the QFIT test to detect people at risk of bowel cancer,^20^ suggesting that our finding has significant predictive sensitivity to be implemented in clinical settings. Additionally, these individuals are already undergoing GI biopsy and so performing an additional IHC stain on the tissue could significantly aid disease-preventative research efforts at a relatively low service cost.

The >80% sensitivity means that if people have *protein misfolding enteropathy* they go on to develop a neurodegenerative disease (median time to diagnosis is 6.9 years, figure 3D). However, the comparatively low specificity means that a negative test cannot reliably identify an individual’s later-life risk. This likely reflects under sampling, for example, that protein misfolding may be occurring in other organs (such as skin, as we have demonstrated previously ^8,21^) that we have not sampled or in a different site within the colon that was not sampled. That led us to investigate the site-specific predominance of these co-pathologies. Our findings demonstrated that the burden of TDP-43 and alpha-synuclein pathology was indeed distinct across our retrospective cohort. TDP-43 pathology was predominantly centred in the left colon (figure 3B) compared to rectal/distal predominance for other proteinopathies such as alpha-synuclein.

Common hypotheses surrounding the involvement of the GI tract in neurodegenerative diseases centre around changes to the global microbiome^22^ and/or physical spreading of aggregates along the vagus nerve, as has been demonstrated for alpha-synuclein.^23,24^ However, the predominating anatomical distribution of alpha-synuclein pathology in our cohort is not within the vagus nerve territory, making physical spread unlikely. Furthermore, the discrete, rather than ubiquitous nature of these pathologies throughout the GI tract makes global changes to the microbiome, again, unlikely contributors to this pathological phenomenon. Rather, we propose that the pathology occurs meta-synchronously across multiple organ systems at the same time, driven by systemic triggers, and cell-type specific susceptibilities that particular organs, such as the brain and the GI tract, share. In line with this, the focus of these pathologies around the splenic flexure and rectum highlights a potential shared underlying mechanism. These sites are both watershed regions, so-called as they lie in the region between two vascular territories. Watershed regions are particularly susceptible to pressure changes and hypoxia, vulnerabilities that have been shown in the brain to confer an increased susceptibility to protein misfolding.^25^ Susceptibility of vulnerable regions, such as watershed regions, could represent a mechanism by which protein misfolding pathology can occur meta-synchronously in response to systemic disease drivers. Correspondingly, we propose that the GI tract represents one such site of end-organ protein misfolding pathology that is readily accessible and that could be used as a window to evaluate this meta-synchronous systemic pathology.

Lastly, we noted a distinct overlap between the proteinopathies within individuals in our cohort. TDP-43 and alpha-synuclein pathologies often co-occurred despite their diverging anatomical distribution in the colon. When we stratified the individuals with PME into groups consisting of either a single, double or triple pathology, we demonstrated that patient survival was decreased in the double and triple pathology groups. The patients included in our cohort would generally be discharged from clinical care after their biopsy was determined to be ‘histologically normal’ – pointing out a new research question, whether a well-tolerated protein clearing intervention could reverse this adverse survival prognosis and prevent or delay neurological disease onset. The perception in the neuroscience field is that these prevention-focused trials should be long term,^26^ however our data suggest that 5-7 years is sufficient to meet survival endpoints for individuals with multi-proteinopathy and for a time-to-event analysis for neurological diagnosis from time of biopsy. Furthermore, as this pathology is readily assessable, we conclude that it could be used as an interim endpoint for target engagement or a treatment response readout for these trials to ensure that therapy is indeed reducing end-organ proteinopathy burden.

In conclusion, detecting PME in routine gastrointestinal biopsies offers a practical, early-access biomarker for neurodegenerative disease. Implementing PME as a predictive tool could transform clinical management by enabling the identification and stratification of at-risk individuals long before central nervous system symptoms emerge. Unlike current interventions, which occur after substantial neuronal loss, early detection of PME would allow pre-emptive therapeutic strategies aimed at preserving neuronal function and delaying phenoconversion. By providing both a predictive and target-engagement biomarker, PME establishes a foundation for prevention-focused clinical trials and opens a new avenue for proactive, rather than reactive, treatment of neurodegenerative disorders.

### Limitations

This study is retrospective and relies on archival gastrointestinal biopsies, which may introduce selection bias and limit control over clinical variables. The cohort represents a single-centre population, and findings may not generalize across diverse populations or age groups. While our staining approach reliably detects pathological protein aggregation and loss-of-function, prospective validation in independent cohorts is required to confirm reproducibility and predictive utility. Despite these limitations, the observed associations between protein misfolding enteropathy, future neurodegenerative disease, and reduced survival provide a robust, hypothesis-generating framework for early detection and prevention-focused clinical trials.

## Data Availability

All non-identifiable data contained in the manuscript. Any other data used in the present study are available upon request to the NHS Grampian Biorepository quoting the tissue request number TR301.

## Contributions

*JMG* conceptualised this study with development contributions from *AJMW, GR, FMW. JMG, GR, AJMW* and *FMW* contributed projected supervision, with *JMG, EM, AJMW* and *GR* providing clinical expertise. *JMG, MP, SM, JI* and *JW* acquired tissue samples *TL, FLR, JMG* and *FMW* acquired histopathological data. Clinical data were curated by *EM, RP* and *GR. FMW, TL* and *JMG* analysed the data. *JMG, FMW* and *TL* wrote the manuscript, which was approved by all the authors.

## Acknowledgements

This work has been supported by a Target ALS Early-Stage ALS Clinician Fellowship (FS-2023-ESC-S2) and an MND Primer award from LifeArc (11.12.2023) to *JMG*; a Target ALS foundation grant to JMG and employing FMW (GCA25107); an NHS Grampian Small Research Grant to *FMW* (SRG 24-16) with co-Is *JMG, GR*, and *EM*. Funders had no role in study design, data collection, data analyses, interpretation, or writing the manuscript. The authors would like to thank the University of Aberdeen Microscopy and Histology Core Facility in the Institute of Medical Sciences, and the NHS Grampian Biorepository staff.

## Data sharing statement

All data with permission for distribution and sharing are included in their entirety within the manuscript. All biopsy material and full clinical data used in this study can be requested for research purposes (anonymised) from the NHS Grampian Biorepository quoting the tissue request number (TR301) – the authors are not custodians of the tissue or data.

